# Differential Benefit of Immune Checkpoint Blockade by *KRAS* Mutation Status in Metastatic Lung Adenocarcinoma: Validation from an Extended Swedish Cohort

**DOI:** 10.1101/2025.04.09.25325505

**Authors:** Ella A. Eklund, Sama I. Sayin, Jonas Smith Jonsson, Hannes van Renswoude, Jan Nyman, Andreas Hallqvist, Clotilde Wiel, Volkan I. Sayin

## Abstract

**Introduction:** Immune checkpoint blockade (ICB) is a standard first-line treatment for advanced-stage non-small cell lung cancer (NSCLC) without actionable oncogenic alterations. *KRAS* mutations, prevalent in 30–40% of Western lung adenocarcinomas, currently lack targeted first-line therapies. This study aimed to assess the predictive value of *KRAS* mutations for clinical outcomes following ICB, validating previous findings in a larger cohort with extended follow-up.

**Methods:** We conducted a retrospective multicenter study including consecutive stage IV NSCLC adenocarcinoma patients (*n* = 424) treated with either ICB or platinum-doublet chemotherapy between 2016–2021 in western Sweden. Patient demographics, tumor characteristics, treatment details, and survival outcomes were retrospectively collected from patient charts and the Swedish National Lung Cancer Registry. *KRAS* mutational status was assessed by next-generation sequencing. Primary endpoints included overall survival (OS) and progression-free survival (PFS), analyzed using Kaplan-Meier curves and multivariate Cox regression.

**Results:** Among 424 patients, 40% harbored *KRAS* mutations (*KRAS*^MUT^). *KRAS*^MUT^ patients showed significant improvement in OS (16 vs. 8 months, *p* < 0.001) and PFS (8 vs. 5 months, *p* < 0.001) with ICB monotherapy compared to chemotherapy. Multivariate analyses confirmed *KRAS* mutation as an independent predictor for better OS (HR 0.533, *p* = 0.018) and PFS (HR 0.523, *p* = 0.018). In contrast, *KRAS* wild-type (*KRAS*^WT^) patients derived no survival advantage from ICB monotherapy. Both *KRAS*^WT^ with *KRAS*^MUT^ populations benefited from chemoimmunotherapy.

**Conclusions:** *KRAS* mutations predict substantial and sustained clinical benefit from first-line ICB monotherapy in advanced NSCLC, whereas *KRAS* wild-type patients do not. Integrating *KRAS* mutation status into clinical practice could guide personalized treatment strategies, optimizing immunotherapy outcomes in advanced-stage lung adenocarcinoma.

## Introduction

Non-small cell lung cancer (NSCLC) remains the leading cause of cancer-related mortality worldwide, with lung adenocarcinoma (LUAD) representing the predominant histological subtype [1]. Recent advancements in molecular profiling and immune checkpoint blockade (ICB) have significantly transformed the therapeutic landscape for patients diagnosed with advanced-stage NSCLC. Currently, ICB—administered either as monotherapy or in combination with chemotherapy or another ICB—constitutes the mainstay first-line treatment option for stage IV NSCLC patients lacking targetable oncogenic alterations [2]. However, only a subset of this patient population responds to ICB treatment [3], and it is crucial to study long-term follow-up outcomes to identify factors predicting treatment response.

Molecular profiling has become a cornerstone of lung cancer diagnostics, guiding therapeutic strategies by identifying actionable genetic alterations. According to current European Society for Medical Oncology (ESMO) guidelines, patients harboring actionable mutations, such as *EGFR* or *ALK* rearrangements, should initially receive targeted tyrosine kinase inhibitors (TKIs) [4]. However, these actionable driver alterations are present only in a minority of NSCLC patients, whereas the majority harbor non-actionable genetic alterations. For the patient subgroup without actionable mutations, guidelines recommend first-line ICB monotherapy for patients with high PD-L1 tumor proportion scores (TPS ≥50%) or chemoimmunotherapy regimens irrespective of PD-L1 expression [2]. Still, response to ICB varies substantially between patients, and currently used predictive biomarkers, such as PD-L1 expression and tumor mutational burden (TMB), remain inconsistent predictors of immunotherapy benefit.

*KRAS* mutations represent the most frequently observed oncogenic mutations in NSCLC, occurring in approximately 30–40% cases among Western populations [5, 6]. The role of *KRAS* mutations, alone or in combination with other risk factors, as prognostic markers for clinical outcomes remains a focus of current research [7-9]. Despite their high prevalence, therapeutic strategies specifically targeting *KRAS*-mutant tumors have only recently begun to emerge, leaving immunotherapy-based regimens as the primary first-line treatment option for these patients [10, 11]. Importantly, *KRAS* mutations have been shown to directly interact with the tumor microenvironment and affect immunogenicity [12-14]. Nevertheless, the prognostic and predictive significance of *KRAS* mutations in the context of real-world outcomes following different ICB-containing treatment regimens remains unclear, and several ongoing trials are evaluating the impact of *KRAS* mutations on response to treatment with ICB [15, 16]. In our previous study, we reported an increased benefit from ICB-containing treatment among patients with *KRAS*-mutated LUAD compared to those without such mutations, whereas ICB monotherapy seemed less effective in *KRAS* wild-type patients [17].

In this retrospective multicenter study, we extend our previous investigation by assembling an expanded cohort comprising all patients diagnosed with stage IV NSCLC who underwent molecular assessment between 2016 and 2021 in Western Sweden. By integrating comprehensive data from the Swedish National Lung Cancer Registry with detailed curation from hospital patient charts, our aim was to assess long-term survival outcomes and elucidate the predictive value of *KRAS* mutational status regarding clinical response to ICB-containing treatments. Furthermore, we sought to validate our earlier observations on the role of *KRAS* mutations in a larger patient cohort with extended follow-up, thereby providing robust, real-world clinical evidence to guide treatment decisions and future biomarker-directed therapeutic strategies.

## Materials and Methods

### Patient population

We conducted a multi-centers retrospective study including all consecutive NSCLC LUAD patients diagnosed with stage IV NSCLC and having molecular assessment performed between 2016-2021 in the Region Västra Götaland (region of West Sweden), Sweden (*n* = 912). The patient receiving any ICB containing treatment or platinum doublet chemotherapy was selected for the study (*n* = 432). For inclusion one treatment cycle was deemed sufficient. Approval from the Swedish Ethical Review Authority (Dnr 2019-04771 and 2021-04987) was obtained prior to study commencement. All patients with available tissue sample where an adenocarcinoma component could not be excluded were systematically assessed with NGS for known genetic drivers (see below) within clinical praxis. Patient demographics (including age, gender, Eastern Cooperative Oncology Group (ECOG) performance status and smoking history), cancer stage, number of metastasis locations, pathological details (PD-L1 grade, histology, mutation status including *KRAS* mutational status and subtype), treatment and outcome data were retrospectively collected from patient charts and the Swedish Lung Cancer Registry.

### Mutational status

Patient selection was done through the pathology regional laboratory information system, where they were referred for molecular assessment. The selected patients underwent next-generation sequencing (NGS) on DNA from FFPE blocks or cytological smears using the Ion AmpliSeq™ Colon and Lung Cancer Panel v2 from Thermo Fisher Scientific until 2019 and thereafter the Thermo Fisher OncomineTM Focus Assay, assessing hotspot mutations in *EGFR, BRAF, KRAS* and *NRAS*. Until June 2017, ALK-fusions were assessed with immunohistochemistry (IHC), and with fluorescence in situ hybridization (FISH) if positive or inconclusive IHC; *ROS1* was analyzed upon request with FISH. Thereafter, *ALK, ROS1* and *RET* fusions were assessed on RNA using the Oncomine Solid Tumor Fusion Panel from Thermo Fisher Scientific. The analyses were done as a part of the standard diagnostic workup process at the Department of Clinical Pathology at Sahlgrenska University Hospital

### ICB containing treatment

During the time period of this study, the most commonly used ICB containing treatments approved for first-line treatment was monotherapy or chemoimmunotherapy. Additionally, anti-CTLA-1 in combination with anti-PD1 therapy was approved in November 2020, but was rarely used in this cohort. The most common monotherapy is Pembrolizumab, a humanized antibody targeting PD-1, for patients with PD-L1high TPS ≥50%. Additionally, Atezolizumab, a humanized antibody targeting PD-L1, is also approved for monotherapy for patients with PD-L1 TPS ≥20%. Chemoimmunotherapy, combined Carboplatin, Pemetrexed, Pembrolizumab, is approved for all patients regardless of PD-L1 TPS.

### Platinum doublet treatment

Platinum doublet treatment (PD) consists of carboplatin or cisplatin in combination with one more non-platinum chemotherapy agent such as pemetrexed, vinorelbine, paclitaxel or gemcitabine.

### Study objectives

The primary outcome of this study was overall survival (OS) and progression free survival (PFS). OS was defined as the time interval between the date of first treatment and the date of death from any cause. PFS was defined as the time interval between the date of first treatment and the date of progression or death whichever came first. Patients alive or lost to follow-up at data were censored at last contact.

### Statistical analysis

Clinical characteristics were summarized using descriptive statistics and evaluated with univariate analysis. Kaplan Meier survival curves were generated to assess OS and PFS. Log-rank test was used to assess significant differences in OS and PFS between groups. Multivariable Cox regression analysis was conducted to compensate for potential confounders. Median follow-up time was calculated using the reverse Kaplan-Meier method. Statistical significance was set at *p* < 0.05 and no adjustments were made for multiple comparisons. Data analysis was conducted using IBM SPSS Statistics version 27 and GraphPad Prism version 9.

## Results

### Patients and tumor characteristics

A total of 432 LUAD patients received ICB-containing treatment or PD chemotherapy in a first line treatment setting. Eight patients were switched to TKI treatment before progression and therefore excluded. A total of 424 patients were included in the final study cohort (Figure 1). Among the included patients, more than 40% harbored a *KRAS* mutation, the majority were female (57%), the median age was 70 years, and a vast majority were current or former smokers (88%) (Table 1). 68% patients had a good PS with ECOG 0-1 at diagnosis (Table 1). The median follow-up time was 67 months. When comparing the baseline characteristics of *KRAS*^WT^ with *KRAS*^MUT^ patients, there were more females, a higher proportion of current and former smokers and slightly higher frequence of patients with PS 0 in the *KRAS*^MUT^ population. We also observed a higher frequency of patients receiving ICB containing treatment (30% vs 40%) and a larger proportion with PD-L1 High in the *KRAS*^MUT^ population (Table 1).

**Table 1.** Patient characteristics of the study population.

| All Stage IV LUAD | Total | KRAS<sup>MUT</sup> | KRAS<sup>WT</sup> |
| --- | --- | --- | --- |
|  | n (%) | n (%) | n (%) |
| Total | 424 (100) | 198 (46.7) | 226 (53.3) |
| Age in years, median (range) | 70 (31-87) | 71 (46-87) | 69 (31-85) |
| Sex |  |  |  |
| Male | 181 (42.7) | 75 (37.9) | 106 (46.9) |
| Female | 243 (57.3) | 123 (62.1) | 120 (53.1) |
| Smoking history |  |  |  |
| Current smoker | 150 (35.4) | 66 (33.3) | 84 (37.2) |
| Former smoker | 223 (52.6) | 122 (61.6) | 101 (44.7) |
| Never smoker | 51 (10.0) | 10 (5.1) | 41 (18.1) |
| Performance status |  |  |  |
| ECOG 0 | 65 (15.3) | 35 (17.7) | 30 (13.3) |
| ECOG 1 | 224 (52.8) | 104 (52.5) | 120 (53.1) |
| ECOG 2 | 107 (25.2) | 51 (25.8) | 56 (24.8) |
| ECOG 3 | 15 (3.5) | 4 (2.0) | 11 (4.9) |
| Missing | 13 (3.1) | 4 (2.0) | 9 (4.0) |
| Histology |  |  |  |
| Adenocarcinoma | 424 (100) | 198 (100) | 226 (100) |
| ICB containing treatment |  |  |  |
| Yes | 149 (35.1) | 79 (39.9) | 70 (31.0) |
| No | 275 (64.9) | 119 (60.1) | 156 (69.0) |
| Treatment |  |  |  |
| Platinum doublet (PD) chemotherapy | 275 (64.9) | 119 (60.1) | 156 (69.0) |
| ICB monotherapy | 75 (17.7) | 44 (22.2) | 31 (13.7) |
| Chemoimmunotherapy | 74 (17.5) | 35 (17.7) | 39 (17.3) |
| Mutation status |  |  |  |
| None known | 186 (43.9) |  | 185 (81.9) |
| KRAS | 198 (46.7) | 198 (100) |  |
| EGFR | 15 (3.5) |  | 15 (6.6) |
| BRAF | 19 (4.5) | 4 (2.0) | 15 (6.6) |
| ALK | 2 (0.5) |  | 2 (0.9) |
| ROS1 | 6 (1.4) |  | 6 (2.7) |
| RET | 3 (0.7) |  | 3 (1.3) |
| PIK3CA | 3 (0.7) |  | 3 (1.3) |
| MET | 3 (0.7) |  | 3 (1.3) |
| HRAS Q61R | 1 (0.2) |  | 1 (0.4) |
| KRAS submutation |  |  |  |
| G12A |  | 12 (6.1) |  |
| G12C |  | 92 (46.5) |  |
| G12D |  | 21 (10.6) |  |
| G12V |  | 33 (16.7) |  |
| G13C |  | 9 (4.5) |  |
| Q61L |  | 9 (4.5) |  |
| Others |  | 22 (11.1) |  |
| PD-L1 grade (%) |  |  |  |
| 0 | 169 (39.9) | 72 (36.4) | 97 (42.9) |
| ≥ 1 | 82 (19.3) | 37 (18.7) | 45 (19.9) |
| ≥ 20 | 46 (10.8) | 15 (7.6) | 31 (13.7) |
| ≥ 50 | 127 (30.0) | 74 (37.4) | 53 (23.5) |
| At last follow up |  |  |  |
| Alive | 46 (10.8) | 24 (12.1) | 22 (9.7) |
| Deceased | 378 (89.2) | 174 (87.9) | 204 (90.3) |
| Survival |  |  |  |
| Median survival (months) | 9.8 | 9.8 | 9.5 |
| No. of metastatic locations at diagnosis |  |  |  |
| 1 | 277 (65.3) | 133 (67.2) | 144 (63.7) |
| 2 | 101 (23.8) | 46 (23.2) | 55 (24.3) |
| 3 | 32 (7.5) | 15 (7.6) | 17 (7.5) |
| >3 | 14 (3.2) | 4 (2.0) | 10 (4.4) |

**Figure 1.**
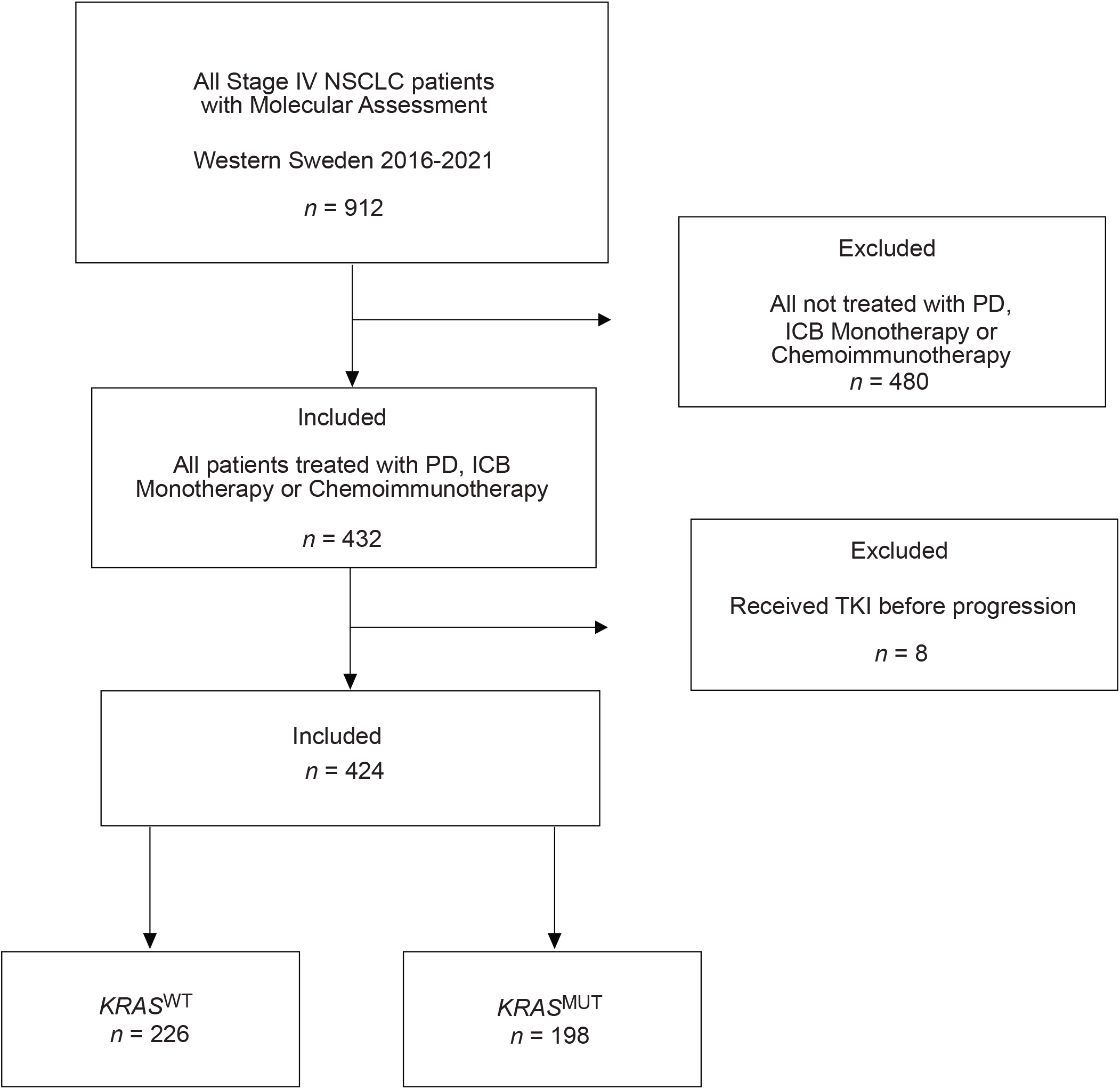
Flowchart showing patient selection for the study.

### ICB containing treatment improves survival outcomes in stage IV LUAD

We initially assessed the efficacy of first line ICB-containing treatment across the entire study cohort *(n* = 424) and there was a significantly improved OS with median 14 vs 8 months (*p* < 0.001) (Supplementary Figure 1A). Additionally, PFS was also significantly longer 7 vs 5 months (*p* < 0.001) (Supplementary Figure 1B). The results were further controlled for confounders with multivariate cox regression. We found that ICB containing treatment was an independent factor for better OS (HR 0.656, 95% CI 0.511-0.843, *p* = 0.006) and PFS (HR 0.613, 95% CI 0.482-0.781, (*p* < 0.001) (Supplementary Figure 1A and 1B).

### Patients with *KRAS* mutations benefit from ICB-containing treatment

Next, we proceeded to evaluate the efficacy of first-line ICB-containing treatment separately for *KRAS*^WT^ and *KRAS*^MUT^ patients. For the *KRAS*^MUT^ group the benefit for OS was 14 vs 8 months (*p* < 0.001). Similarly, in the *KRAS*^WT^ group there was an OS benefit with 15 vs 8 months (*p* = 0.032) (Figure 2A). However, when results were further controlled for confounders with multivariate cox regression, we found that ICB-containing treatment was an independent factor for better OS (HR 0.562, 95% CI 0.387-0.816, (*p* = 0.002) in the *KRAS*^MUT^ group, while it was not an independent factor for better OS in *KRAS*^WT^ (HR 0.716, 95% CI 0.507-0.1013, *p* = 0.059) (Figure 2A).

**Figure 2.**
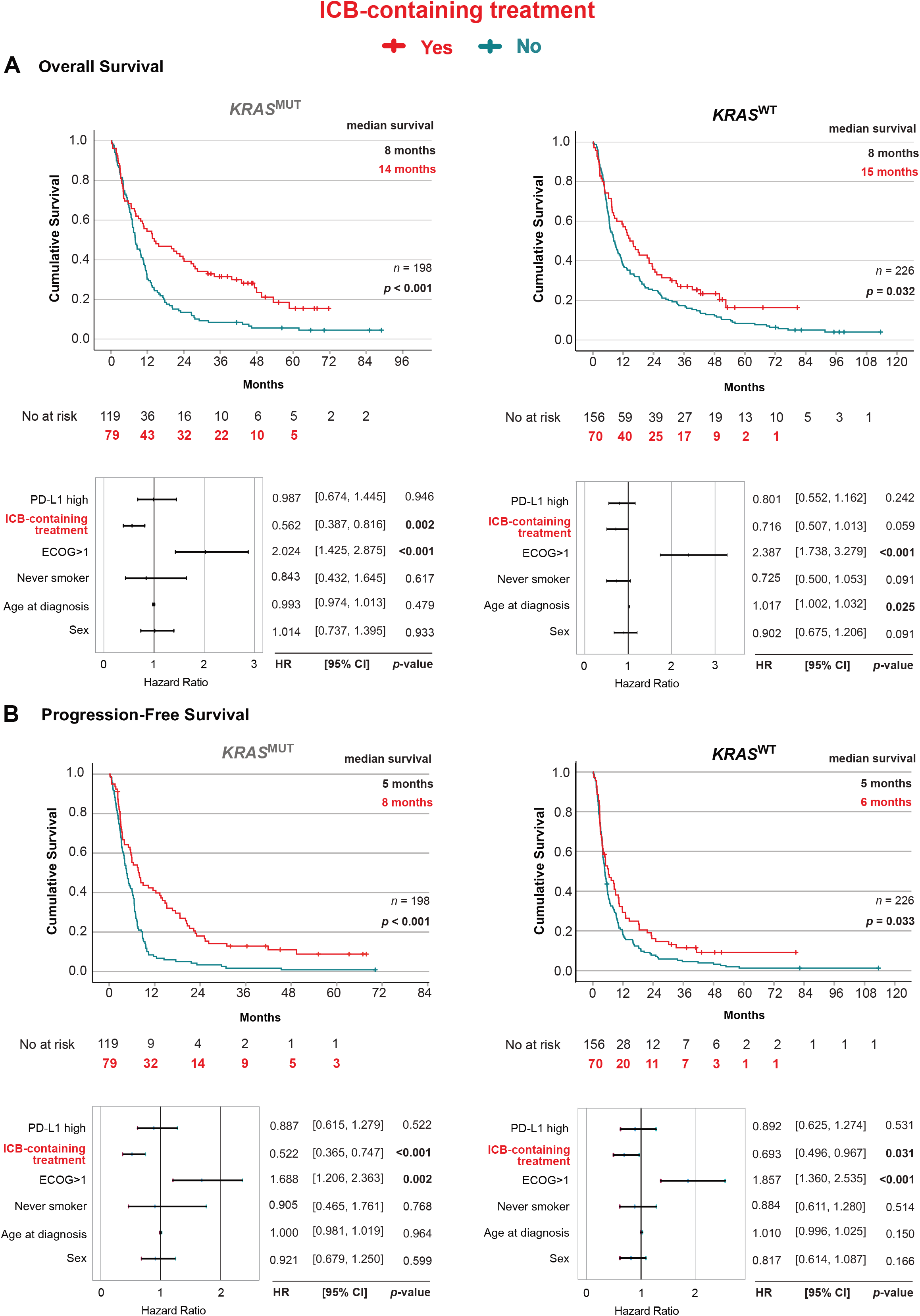
Impact of ICB-containing treatment on (**A)** overall survival and (**B)** progression-free survival in LUAD subgroups. Top Panels: Kaplan-Meier estimates comparing survival outcomes between patients who received ICB-containing treatment (Yes, Red) or PD alone (No, Teal). Bottom Panels: Forest plot of multivariable COX regression analysis within LUAD subgroups. Left top and bottom panels: *KRAS* mutation (*KRAS*^MUT^), Right top and bottom panels: *KRAS* wildtype (*KRAS*^WT^).

*KRAS*^MUT^ group showed a larger benefit in PFS, 8 vs 5 months (*p* <0.001), than in *KRAS*^WT^ group with 6 vs 5 months (*p* = 0.033) (Figure 2B). When further controlled for confounders with multivariate cox regression it confirmed ICB containing treatment as an independent factor for better PFS both for *KRAS*^MUT^ (HR 0.522, 95% CI 0.365-0.747, *p* <0.001) and *KRAS*^WT^ (HR 0.693, 95% CI 0.496-0.967, *p* = 0.031) (Figure 2B).

### ICB monotherapy improves survival outcomes in *KRAS*^MUT^ but not in *KRAS*^WT^

When assessing ICB monotherapy, we found that *KRAS*^MUT^ group clearly benefitted with 16 vs 8 months (*p* < 0.001), whereas *KRAS*^WT^ group did not benefit compared to PD treatment, with OS 8 vs 8 months (*p* = 0.648) (Figure 3A). In multivariate analysis receiving ICB mono was independent factor for better OS in the *KRAS*^MUT^ group (HR 0.533, 95% CI 0.311-0.912, *p* = 0.018) (Figure 3A).

**Figure 3.**
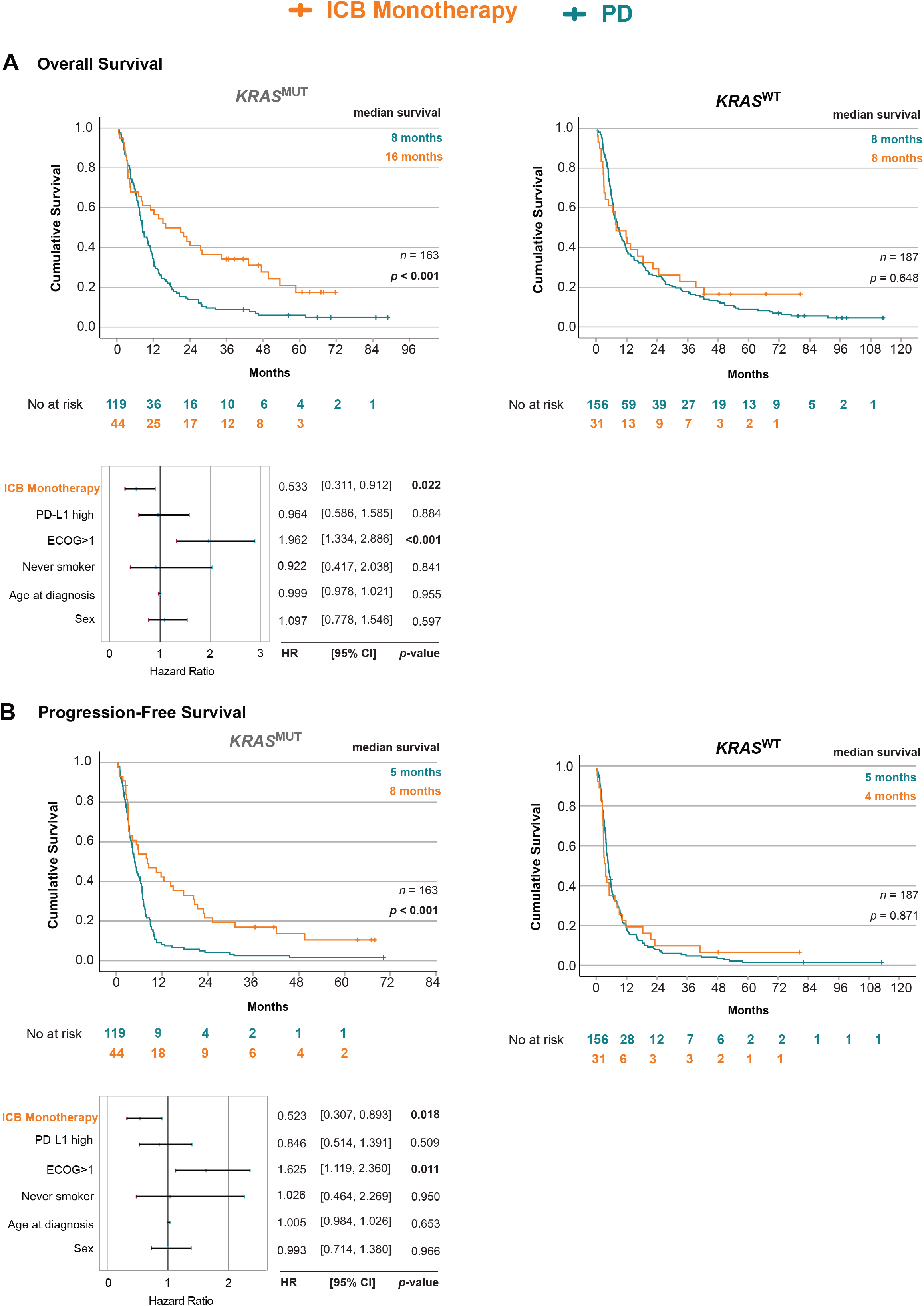
Impact of ICB monotherapy on (**A)** overall survival and (**B)** progression-free survival in LUAD subgroups. Top Panels: Kaplan-Meier estimates survival outcomes between patients who received ICB monotherapy (Yes, Orange) or PD alone (No, Teal). Bottom Panels: Forest plot of multivariable COX regression analysis within LUAD subgroups. Left top and bottom panels: *KRAS* mutation (*KRAS*^MUT^), Right top panel: *KRAS* wildtype (*KRAS*^WT^).

*KRAS*^MUT^ group clearly benefitted in PFS from ICB monotherapy, with 8 vs 5 months (*p* < 0.001) compared to PD. However, the PFS analysis did not show any benefit of ICB monotherapy for the *KRAS*^WT^ group with 4 vs 5 months (*p* = 0.871) (Figure 3B). In multivariate analysis, receiving ICB monotherapy was independent factor for better PFS in the *KRAS*^MUT^ group (HR 0.523, 95% CI 0.307-0.893, *p* = 0.018) (Figure 3B).

### Both patients with and without *KRAS* mutations benefit from chemoimmunotherapy treatment

We further assessed chemoimmunotherapy compared to PD treatment. *KRAS*^MUT^ patients had an improved OS on chemoimmunotherapy compared to PD, 14 vs 8 months (*p* = 0.009) and *KRAS*^WT^ had OS 21 vs 8 months (*p* = 0.011) (Figure 4A). There was also a benefit in PFS for *KRAS*^MUT^ with 8 vs 5 months (*p* < 0.001), and 9 vs 5 months for *KRAS*^WT^ (*p* = 0.006) (Figure 4B). When further controlled for confounders with multivariate cox regression, chemoimmunotherapy was an independent factor for better OS and PFS for both the *KRAS*^MUT^ and *KRAS*^WT^ groups (Figure 4A, B).

**Figure 4.**
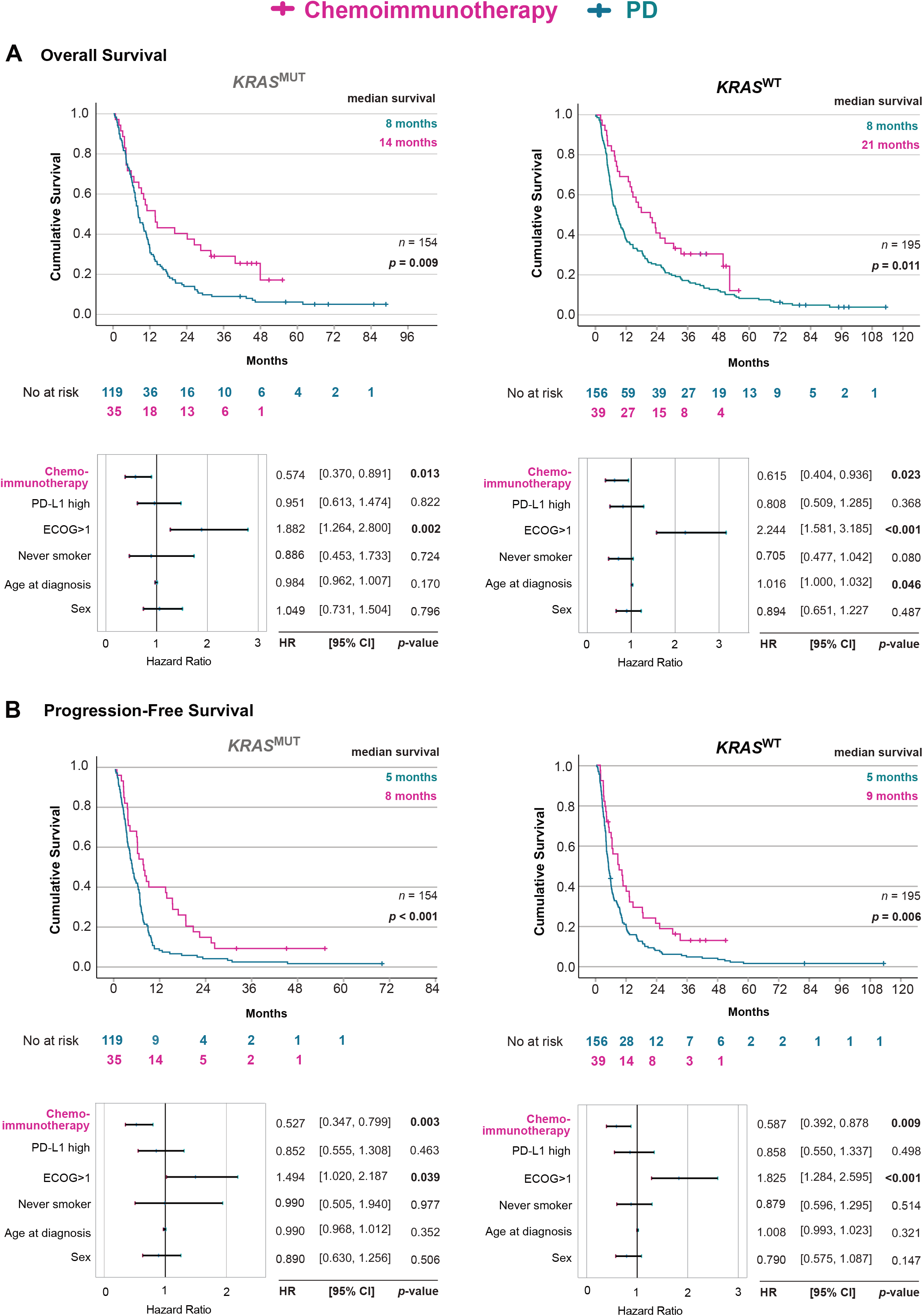
Impact of chemoimmunotherapy on (**A)** overall survival and (**B)** progression-free survival in LUAD subgroups. Top Panels: Kaplan-Meier estimates comparing survival outcomes between patients who received Chemoimmunotherapy (Yes, Magenta) or PD alone (No, Teal). Bottom Panels: Forest plot of multivariable COX regression analysis within LUAD subgroups. Left top and bottom panels: *KRAS* mutation (*KRAS*^MUT^), Right top and bottom panels: *KRAS* wildtype (*KRAS*^WT^).

## Discussion

In this extended retrospective, multicenter analysis of patients diagnosed with stage IV lung adenocarcinoma treated with ICB, we validated and expanded our previous findings by highlighting the crucial predictive role of *KRAS* mutations in response to immunotherapy. Our results underscore that *KRAS*^MUT^ patients achieve significant improvement in both OS and PFS following ICB monotherapy compared to platinum doublet chemotherapy alone. In contrast, *KRAS*^WT^ patients showed no significant benefit from monotherapy.

This differential response is particularly notable given the historical perception of *KRAS* mutations as a negative prognostic indicator in NSCLC. Recent literature increasingly supports the idea that *KRAS* mutations may be prognostic for improved outcomes in NSCLC patients treated with ICB [18]. A meta-analysis by Zhao et al. (2024) found that *KRAS* mutation has beneficial effects on OS and PFS in NSCLC patients treated with immunotherapy [19]. Similar to our findings, Li et al. (2022) reported that following immunotherapy, the OS and PFS of patients with *KRAS* mutations were significantly higher than those in the non-*KRAS* mutant group [20].

Studies have shown that *KRAS* mutations in NSCLC are associated with a distinct tumor microenvironment characterized by increased immunogenicity and inflammation, potentially enhancing sensitivity to immune checkpoint inhibitors [21]. Our findings align closely with those of Liu et al. (2020), who demonstrated superior efficacy of anti-PD-1/PD-L1 therapies among *KRAS*-mutant patients associated with heightened immunogenic signatures [12]. Further supporting our findings, Torralvo et al. (2019) also observed improved clinical outcomes following immunotherapy in *KRAS*-mutant populations [22].

In contrast, Noordhof et al. (2021) found no difference in survival depending on *KRAS* mutation after ICB monotherapy [23]. Mok et al. showed that in KEYNOTE-042, the clinical benefit from pembrozliumab was maintained regardless of KRAS status compared to chemotherapy [24]. However, unlike us, these above studies have included NSCLC patients with all histological subtypes, and only patients with PD-L1 >50%. Given PD-L1 has been shown to be unreliable biomarker of response to immunotherapy [25], our cohort removes this filter to include a more representative population for judging biological underpinnings of *KRAS*-mutated LUAD response to immunotherapy. Additionally, our real-world cohort study provides enhanced robustness due to significantly extended follow-up, reinforcing *KRAS* mutations as positive predictive biomarkers for ICB treatment efficacy.

Nevertheless, the response within *KRAS*-mutant NSCLC is not universally uniform. Recent studies have highlighted that specific *KRAS* mutation subtypes might have distinct clinical behaviors and potentially alter responsiveness to immunotherapy [20, 26]. In contrast, Urtecho et al. (2025) found no differences in response to ICB between *KRAS* subtype groups [27].

Although our analysis did not differentiate *KRAS* subtypes, such granularity may further improve patient stratification and individualized therapeutic selection in future studies. Additionally, emerging evidence indicates significant heterogeneity related to co-occurring genetic alterations, such as *STK11, KEAP1, TP53* and *LRP1B* [7, 28]. Although our study did not comprehensively evaluate co-mutations, this aspect remains critical for future investigations and biomarker refinement.

The strengths of our study include a large, well-characterized, and regionally representative patient cohort with comprehensive long-term follow-up data, alongside the utilization of standardized molecular testing practices aligned with routine clinical care. Limitations include the retrospective nature, potential selection bias, and absence of detailed co-mutation or *KRAS*-subtype analyses. These limitations indicate important directions for subsequent prospective validation studies.

In conclusion, our findings clearly demonstrate that *KRAS* mutation status significantly predicts long-term benefit from ICB monotherapy in advanced NSCLC. Our findings notably caution against the use of ICB monotherapy in patients without *KRAS* mutations. Specifically, *KRAS*-mutant patients gain substantial and durable survival improvements, whereas *KRAS* wild-type patients appear to derive limited or no benefit from immunotherapy without concurrent chemotherapy. Integrating *KRAS* mutational analysis into clinical decision-making may thus enable more precise, personalized treatment strategies, optimizing immunotherapy outcomes for patients with advanced-stage NSCLC.

## List of abbreviations

ECOG: Eastern Cooperative Oncology Group
ICB: Immune Checkpoint Blockade
NSCLC: Non-Small Cell Lung Cancer
NGS: Next Generation Sequencing
PD: Platinum Doublet
PD-1: Programmed Cell Death 1
PD-L1: Programmed Death Ligand 1
PS: Performance Status
OS: Overall Survival
TPS: Tumor Proportion Score

## Ethical Statement

Approval from the Swedish Ethical Review Authority (Dnr 2019-04771 and 2021-04987) was obtained prior to the commencement of the study. No informed consent was required due to all data being presented in a de-identified form.

## Consent for publication

Not applicable. Patient consent statements were not required due to the retrospective nature of this study.

## CRediT Author contributions

**EAE, SIS**: Conceptualization; Formal analysis; Investigation; Methodology; Supervision; Validation; Visualization; Writing - original draft; and Writing - review & editing

**JSJ** and **HR**: Data curation; Formal analysis

**JN, AH** Conceptualization; Supervision; Writing - review & editing

**CW, VIS:** Conceptualization; Project administration; Supervision; Funding acquisition; Writing - review & editing

## Data availability

The datasets used and/or analyzed during the current study is available from the corresponding author on reasonable request.

## Declaration of generative AI and AI-assisted technologies in the writing process

During the preparation of this work the authors used ChatGPT4o to proof text and improve language for clarity. After using this tool, the authors reviewed and edited the content as needed and take full responsibility for the content of the publication.

## List of Supplemental material

Supplemental Figure 1. PDF

**Supplementary Figure 1.**
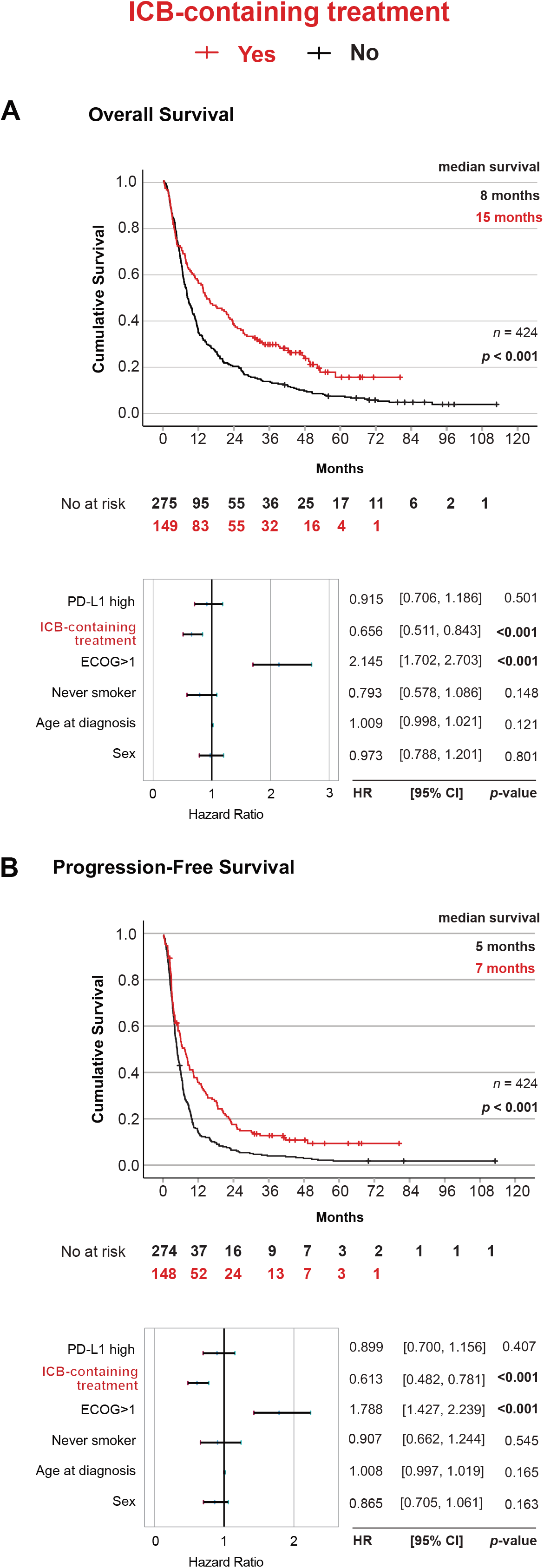
Impact of ICB-containing treatment on (**A)** overall survival and (**B)** progression-free survival in all LUAD patients. Top Panels: Kaplan-Meier estimates comparing survival outcomes between patients who received ICB-containing treatment (Yes, Red) or PD alone (No, Teal). Bottom Panels: Forest plot of multivariable COX regression analysis among all LUAD patients.

